# Who dies from COVID-19? Post-hoc explanations of mortality prediction models using coalitional game theory, surrogate trees, and partial dependence plots

**DOI:** 10.1101/2020.06.07.20124933

**Authors:** Russell Yang

## Abstract

As of early June, 2020, approximately 7 million COVID-19 cases and 400,000 deaths have been reported. This paper examines four demographic and clinical factors (age, time to hospital, presence of chronic disease, and sex) and utilizes Shapley values from coalitional game theory and machine learning to evaluate their relative importance in predicting COVID-19 mortality. The analyses suggest that out of the 4 factors studied, age is the most important in predicting COVID-19 mortality, followed by time to hospital. Sex and presence of chronic disease were both found to be relatively unimportant, and the two global interpretation techniques differed in ranking them. Additionally, this paper creates partial dependence plots to determine and visualize the marginal effect of each factor on COVID-19 mortality and demonstrates how local interpretation of COVID-19 mortality prediction can be applicable in a clinical setting. Lastly, this paper derives clinically applicable decision rules about mortality probabilities through a parsimonious 3-split surrogate tree, demonstrating that high-accuracy COVID-19 mortality prediction can be achieved with simple, interpretable models.

## Introduction

Interpretable machine learning is critically important in healthcare, and clinicians seek explanations that justify and rationalize model predictions [1]. Medical professionals also prefer parsimonious machine learning methods because of their explainability and because they are more likely to conform to operational guidelines, which often include fixed attribute scores [2]. Thus, feature extraction is often eschewed in medical research because it reduces interpretability [2].

The incubation period of COVID-19 is about 5.2 days [3], and there is a median length of 14 days between onset of symptoms and death [4]. COVID-19 symptoms include pneumonia, fever, fatigue, and dry cough [5], and risk factors include pre-existing health conditions (asthma, chronic lung/kidney disease, diabetes, hemoglobin disorders, being immunocompromised, liver/heart disease), old age, and obesity [6]. COVID-19 mortality also varies among different ethnicities, potentially due to discrimination, economic disadvantages, unequal access to health care, and other factors [7].

ICU resources are scarce and ethical dilemmas arise in deciding how to allocate limited hospital resources [8]. The demand for ICUs and beds in hospitals is increasing as the number of cases rise, and ICUs already had high occupancy before the pandemic. Previous estimates of mean hourly occupancy of ICUs put the number at about 68.2% [9].

Much of the current COVID-19 informatics literature focuses on macro-level disease forecasting using machine learning and statistical techniques, with few studies focusing on individual-level predictions. For example, [10] utilizes a SEIR (Susceptible-Exposed-Infectious-Removed) differential equation-based model to predict the sizes and peaks of the COVID-19 pandemic, and [11] utilizes a logistic model to understand the COVID-19 case trend. One study published in Nature Machine Intelligence used various biomarkers (lactic dehydrogenase, lymphocyte and high-sensitivity C-reactive protein) to achieve advanced individual-level COVID-19 mortality predictions with 90% accuracy [12]. We hypothesize that demographic and temporal risk factors can explain COVID-19 mortality as well, avoiding the time and cost associated with biomarker measurement.

Recently, epidemiological datasets with demographic, geographic, and temporal data have become available and have opened up new dimensions for COVID-19 modeling. One such dataset is [13]. This study focuses on ranking the relative importance of age, time to hospital after symptom onset, sex, and presence of chronic disease in COVID-19 mortality prediction and developing a framework for local interpretation of COVID-19 mortality predictions in clinical settings.

## Methods

### Sourcing and Preprocessing

This analysis utilized publicly available individual-level epidemiological data as of June 4th, 2020 [13]. The dataset includes various temporal, demographic, geographic, and environmental attributes, including age, sex, city, province, country, sourced from Wuhan or elsewhere, latitude, longitude, etc. It was aggregated from various sources and is extremely sparse. Several preprocessing steps were employed to filter and clean the data.

4 suspected risk factors were studied as explanatory variables: age, time from onset of symptoms to hospital admission, sex, and presence of chronic disease. The outcome variable was binary: either recovery or mortality. The dataset was subsetted to include only relevant columns. The sex binary categorical variable was encoded to numeric values. Samples were removed from the analysis if they had missing values for any of the relevant variables. There was heterogeneity in clinical variable annotation, so various values of outcome (‘discharge’, ‘discharged’, ‘Discharged’, ‘recovered’) were coded to 0 (recovery) and other values (‘died’, ‘death’) were coded to 1 (mortality). Patients with other outcome values (‘severe’, ‘stable,’ ‘Symptoms only improved with cough. Currently hospitalized for follow-up.’) were removed from the analysis. For samples where an age range was given instead of a single number, the lower and upper limits of the range were averaged to produce a single number. One sample was assumed to have a coding error in the date_onset_symptoms column and was removed. A new derived column to represent time from onset of symptoms to hospital admission was created (time_to_hospital = date_admission_hospital - date_onset_symptoms). One sample had a negative value for time_to_hospital, which was assumed to be the result of a coding error and was removed.

After filtering and cleaning the dataset, 184 viable patients remained. These 184 patients may not necessarily be representative of the global population (in terms of geographic location, healthcare quality, etc.) because many samples had to be discarded in the preprocessing steps; nonetheless, we hope that the relative importance of age, sex, time to hospital, and presence of chronic disease will be relatively consistent between this sample and the global population. Furthermore, some individuals may have experienced mortality after being discharged from the hospital, but that information was not included in the dataset. Here, we provide visualizations and descriptive statistics to understand the 184-patient dataset. Fig 1 provides histograms of the continuous covariates and Table 1 provides summary statistics for the dataset. As shown in Table 1, the mean age of patients was about 48.02 (SD 18.62). 63.59% of patients were male. Chronic disease was present in 20.11% of individuals, and the average time to hospital was 5.17 (SD 4.28). Approximately 25.54% of individuals in the dataset experienced mortality.

**Fig 1:**
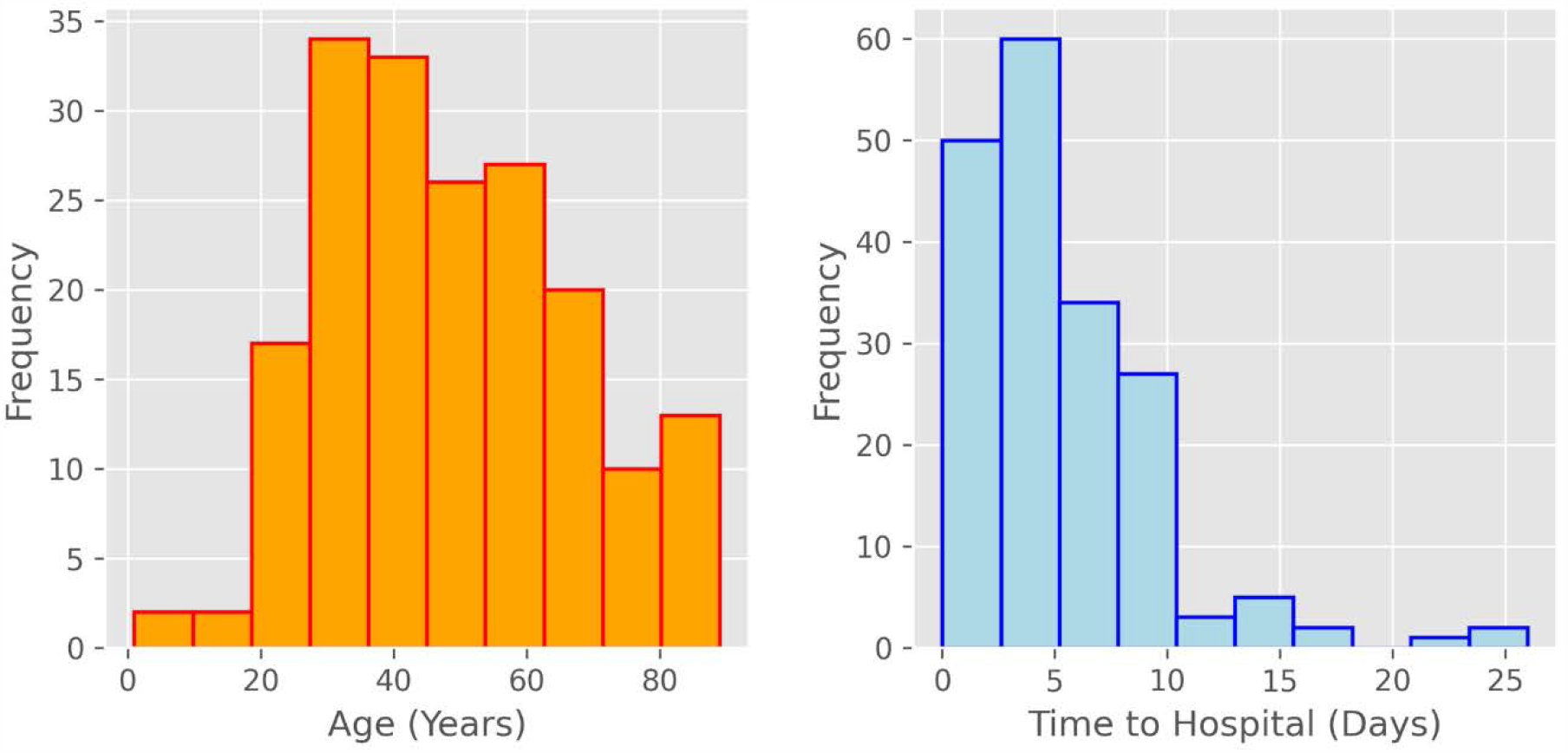
Histograms for the two continuous covariates (age and time_to_hospital)

**Table 1.** Descriptive statistics for variables in the 184-patient dataset.

|  | age (yrs) | time_to_hospital (days) | sex | chronic_disease_binary | outcome |
| --- | --- | --- | --- | --- | --- |
| **mean** | 48.019022 | 5.168478 | 0.635870 | 0.201087 | 0.255435 |
| **std** | 18.615785 | 4.279687 | Not applicable for binary data |  |  |
| **min** | 1 | 0 |  |  |  |
| **Q1** | 33 | 2 |  |  |  |
| **median** | 46 | 5 |  |  |  |
| **Q3** | 61 | 7 |  |  |  |
| **max** | 89 | 26 |  |  |  |

An XGBoost model was trained for binary classification of patient mortality/recovery. XGBoost utilizes a gradient tree boosting algorithm and provides state-of-the-art classification performance in many scenarios [14]. The algorithm is highly scalable and utilizes minimal machine resources [14]. The model was trained with default parameters using the Python xgboost package. Table 2 shows various classification metrics of the XGBoost model when it was trained on 70% of the data and tested on the remaining 30%. The model achieves an testing accuracy of 0.91.

**Table 2:** Classification report for XGBoost model predictions on test set.

|  | Precision | Recall | F1 Score | Support |
| --- | --- | --- | --- | --- |
| **0 (Recovery)** | 0.95 | 0.93 | 0.94 | 44 |
| **1 (Mortality)** | 0.77 | 0.83 | 0.80 | 12 |
| **Accuracy** |  |  | 0.91 | 56 |
| **Macro Avg** | 0.86 | 0.88 | 0.87 | 56 |
| **Weighted Avg** | 0.91 | 0.91 | 0.91 | 56 |

### Shapley Additive Explanations (SHAP)

SHAP is a method for model interpretation that relies on the Shapley value, a solution concept in coalitional game theory. In coalitional game theory, the Shapley value represents a distribution of a collective payoff/prediction among multiple participants/features. In feature interpretation using Shapley values, predictions are compared between models with and without each feature so that importance values can be assigned to each feature. Shapley values are given by the following formula, where F is the feature set, the summation is over all the possible feature subsets, the expression in brackets is the difference in predictions between a model trained on the feature subset and a model trained on the same feature subset but also with feature i, and the fraction is a factor for averaging [15]:

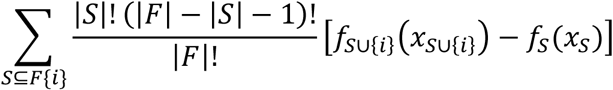

Intuitively the Shapley value can be interpreted as the expected value of the marginal contribution to the coalition, and it is computed by adding each feature to a model and understanding how it impacts the prediction. Shapley feature attribution methods possess several desirable properties, including local accuracy, missingness, and consistency [15]. The method used in this paper is Tree SHAP, which is a variant of SHAP for decision tree models. Tree SHAP improves the time complexity of SHAP from exponential to polynomial [16].

### Skater

The Skater package was also employed for model interpretation. The package was used to create model-agnostic partial dependence plots and perform local interpretation using LIME (Local Interpretable Model-Agnostic Explanations). Additionally, parsimonious tree surrogates were created. Partial dependence plots specify the marginal effect of features on the response variable in a model. According to [17], the partial dependence is given by the following formula, where S is a subset of predictor indices and C is the complement of S:

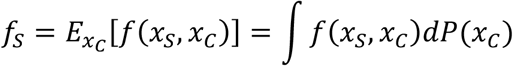

In practice, partial dependence is estimated using the following formula, where N is the number of samples in the training set and *x*_*C*1_ through *x*_*CN*_ are observed values of *x*_*C*_ from the training set [17]:

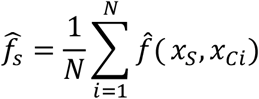

LIME is a technique that uses local approximations to a machine learning model to provide interpretations of the prediction of any sample [18]. Roughly speaking, LIME perturbs the model many times to determine the influence of each explanatory variable on the outcome variable. LIME allows for rapid and clinically useful local interpretation of the model’s predictions. Furthermore, LIME explanations are locally faithful [18]. Surrogate trees are approximations of complex models (such as those produced by the XGBoost algorithm). They are model-agnostic since they can be trained by observing inputs and outputs of the underlying model [19]. Unfortunately (but unsurprisingly), a tradeoff exists between fidelity (how well the surrogate can approximate the original model) and model complexity [19].

## Results

### Shapley Additive Explanations

A TreeExplainer from the shap package in Python was used to calculate Shapley values. The TreeExplainer object can be used for global interpretations of the model as well as local interpretations of the prediction for any individual. In Fig 2, the relative importance of explanatory variables is plotted. According to the Shapley values, age is the most important of the 4 features, followed by time_to_hospital, chronic_disease_binary, then sex.

**Fig 2:**
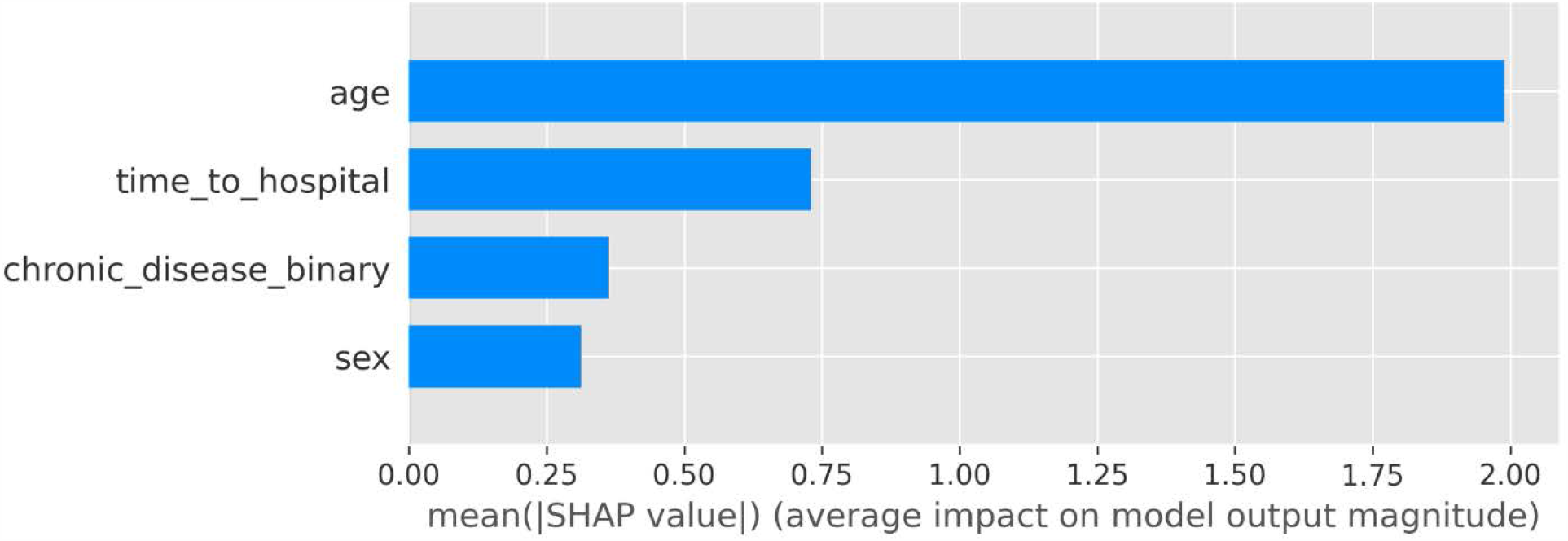
Barplot of relative feature importance of explanatory variables as assessed by mean absolute value of Shapley value.

Fig 3 shows example local interpretations for two patients. In the figure, values of certain features ‘push’ the prediction from an initial base value (bias) to a final model output value. In the first patient, the low age (38) was the major factor that pushed the patient towards a smaller model output value, whereas in the second patient, the high age (82) pushed the patient towards a higher value. Also, being male pushed the model output up in the first patient and being female pushed the model output down in the second patient. In the first individual, absence of chronic disease pushes the model output down, while presence of chronic disease pushes the output up in the second individual. Interestingly, a time to hospital value of 7 pushes one individual down and the other up.

**Fig 3:**
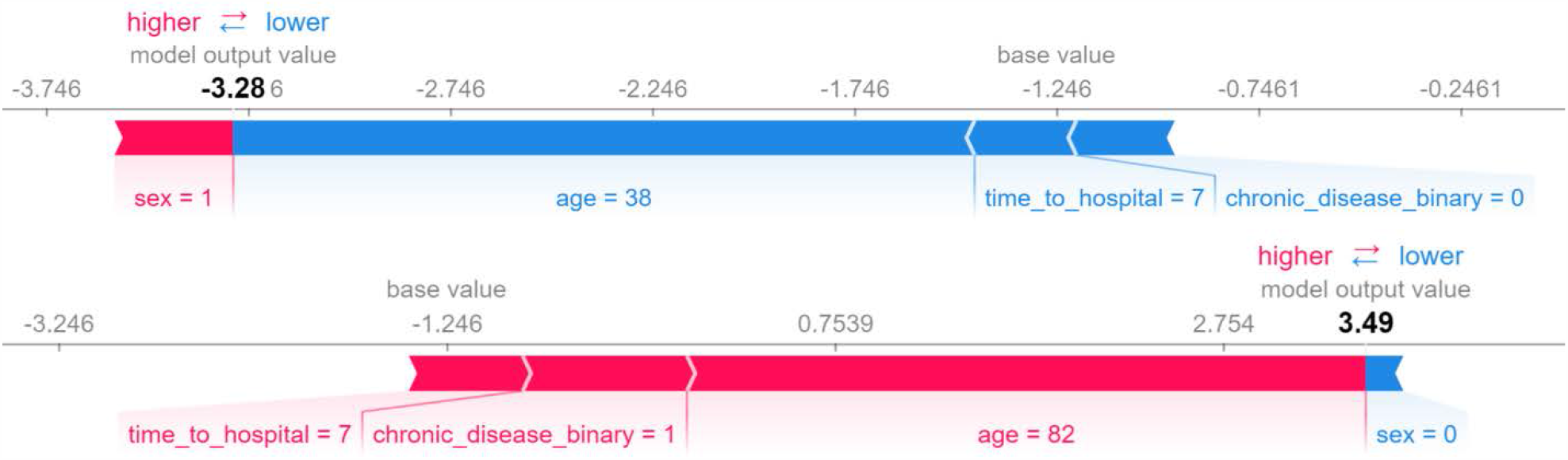
Sample local explanations for a negative and positive individual.

Fig 4, created using the shap package, shows local interpretations for all patients on one graph. The magnitude of the SHAP value quantifies the importance of the feature in the model, and each dot signifies a Shapley value for an individual’s feature.

**Fig 4:**
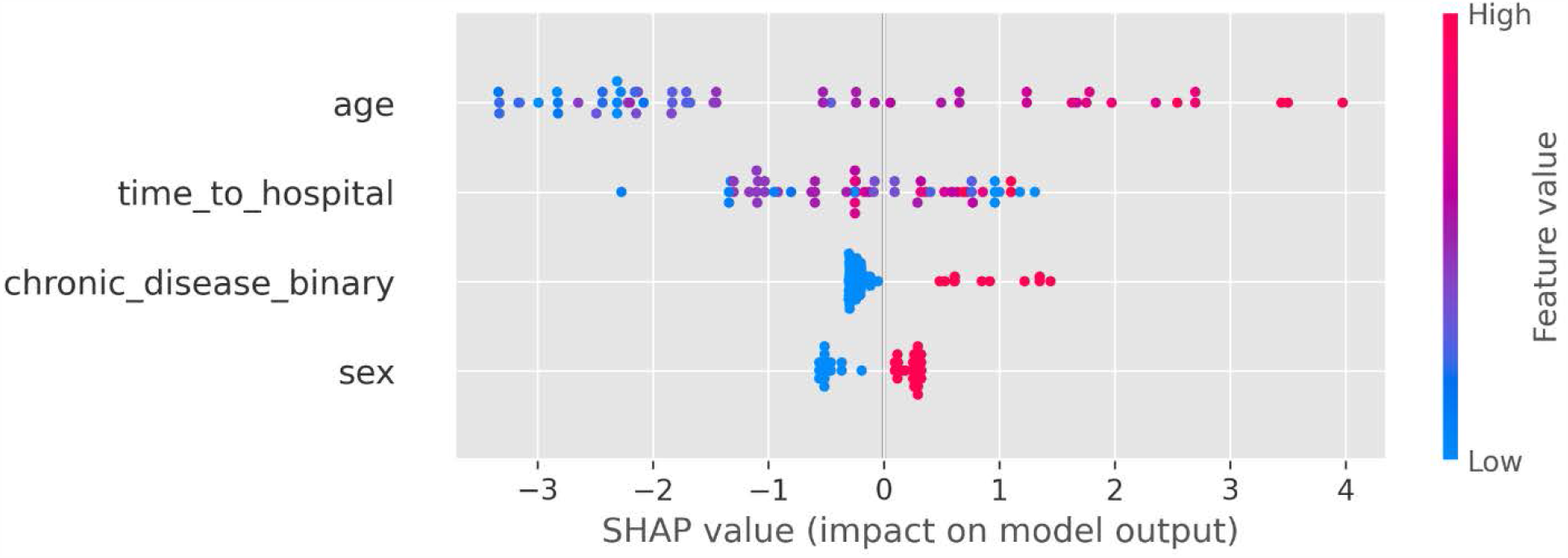
SHAP Interpretation for all patients.

Partial dependence plots were created for each of the four explanatory variables (Fig 5). Higher values of age are associated with higher SHAP values. Values of 1 for sex (male) are associated with higher SHAP values than 0 for sex (female). Likewise, values of 1 for chronic_disease_binary (chronic disease present) are associated with higher SHAP values than 0 for chronic_disease_binary (chronic disease absent). The partial dependence plot for time_to_hospital exhibits heteroskedasticity and cannot be easily interpreted.

**Fig 5:**
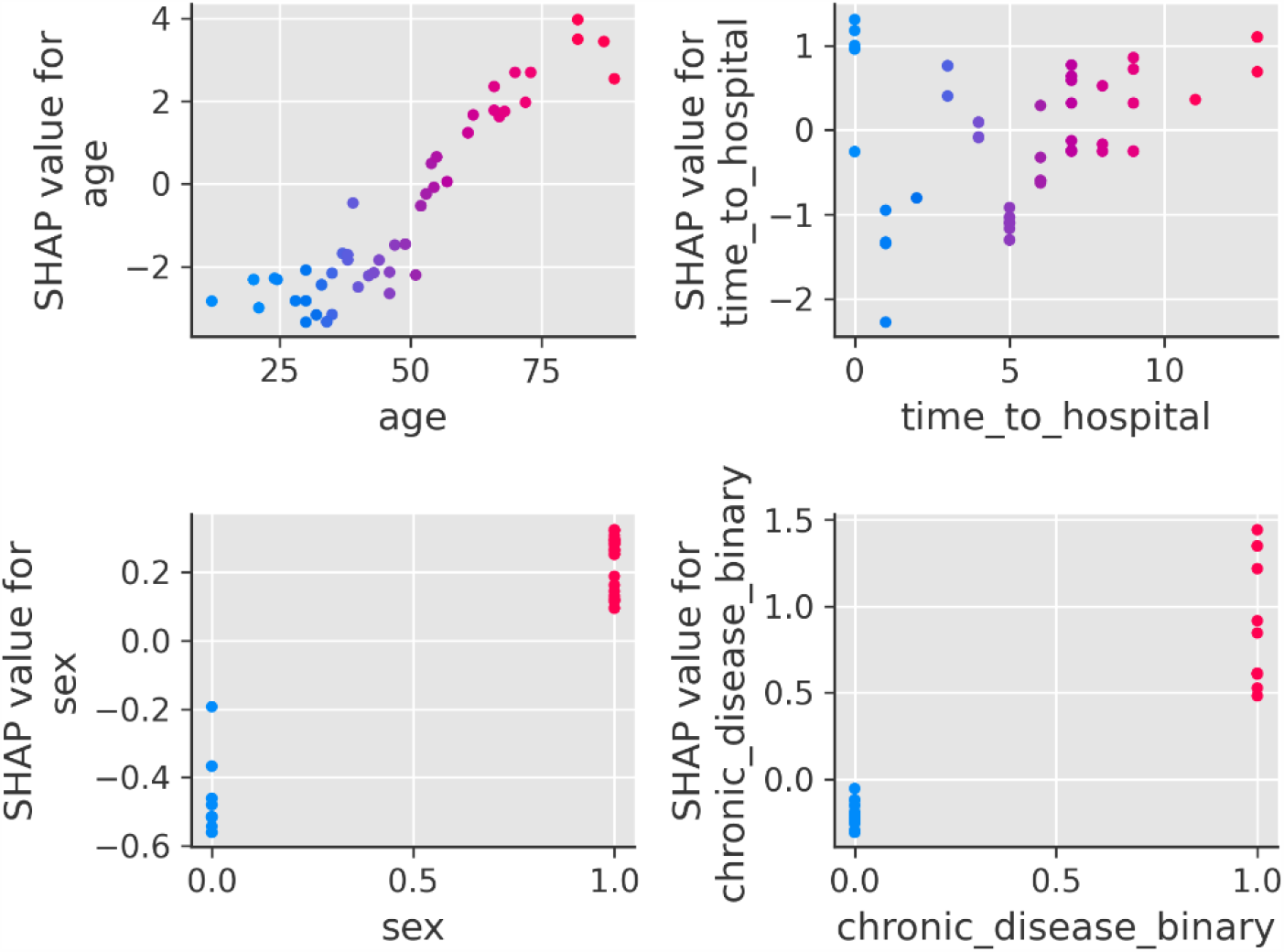
Partial dependence plots for each of the 4 explanatory variables.

Fig 6 shows the partial dependence plot for age, and points are colored by time_to_hospital to elucidate potential interactions between age and time_to_hospital.

**Fig 6:**
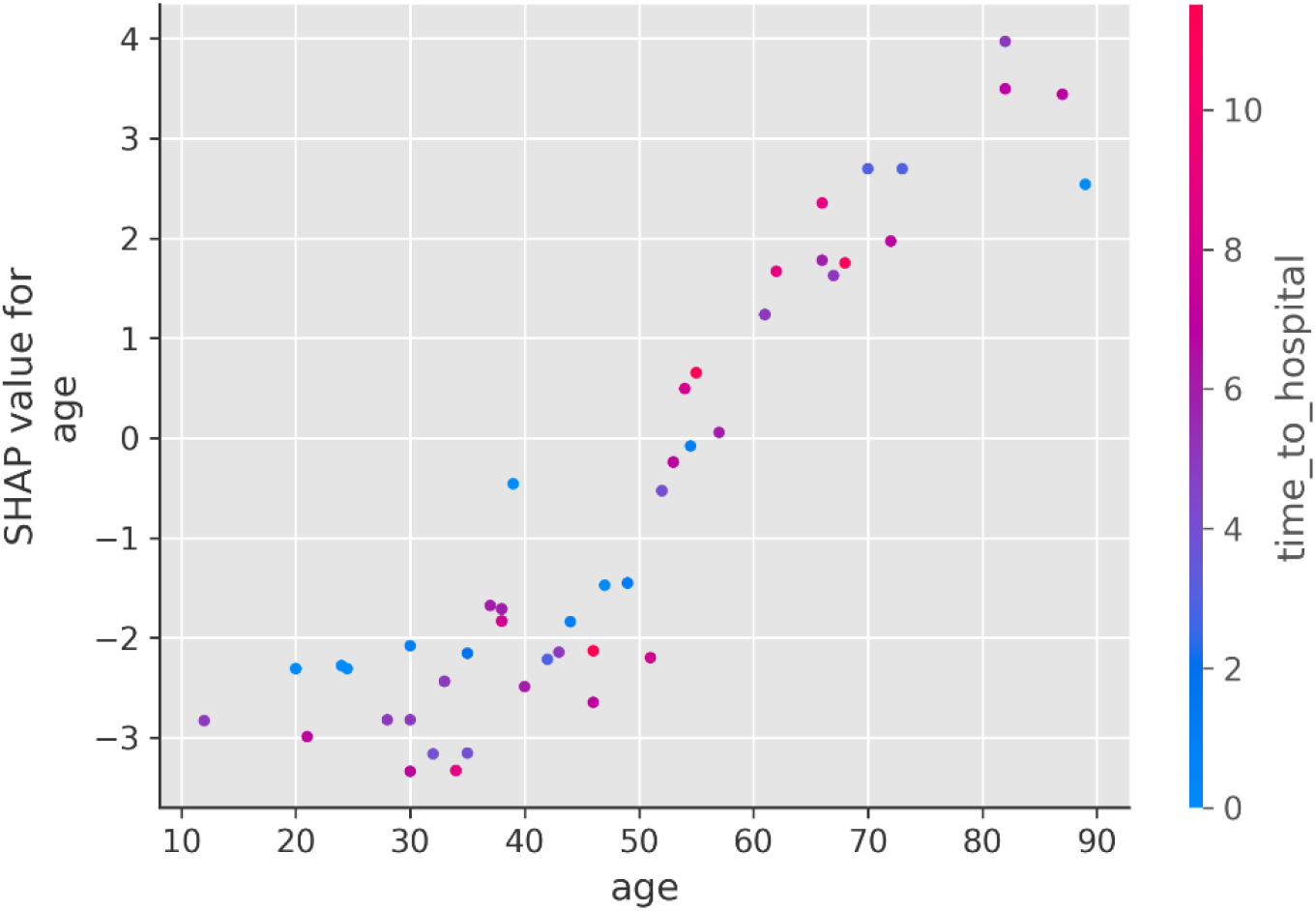
Partial dependence plot for age with interaction index set to time_to_hospital.

### Skater Interpretations

The skater package in Python was also used to perform interpretation analyses. Skater, like shap, has global and local interpretation abilities. As shown in Fig 7, the skater packages provides a similar ordering of feature importance as the shap package. Age is the most important feature by far, followed by time_to_hospital. However, skater ranks sex as more important than chronic_disease_binary, while shap ranks chronic_disease_binary as more important than sex.

**Fig 7:**
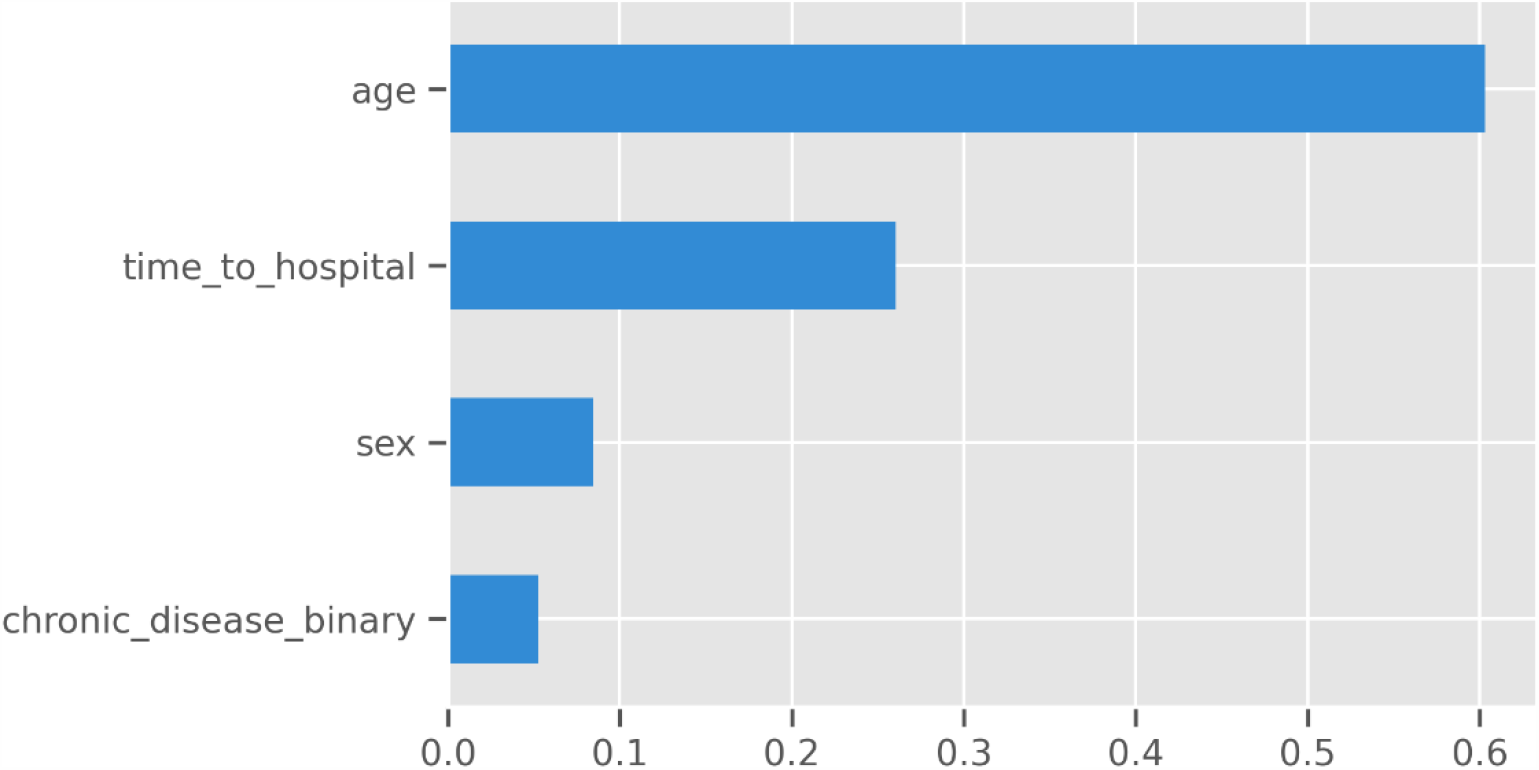
Barplot of relative feature importance of explanatory variables as assessed by skater package.

A LimeTabularExplainer object was then created using the skater package. LIME (Local Interpretable Model-Agnostic Explanations) was used to perform local interpretations. Fig 8 lists the factors contributing to recovery/death and summarizes them in a table, where orange colored factors are those that contribute to mortality and blue colored factors are those that contribute to recovery. For example, in the bottom patient (predicted to experience mortality), the high age, presence of chronic disease, and time to hospital all contribute to the high probability of death.

**Fig 8:**
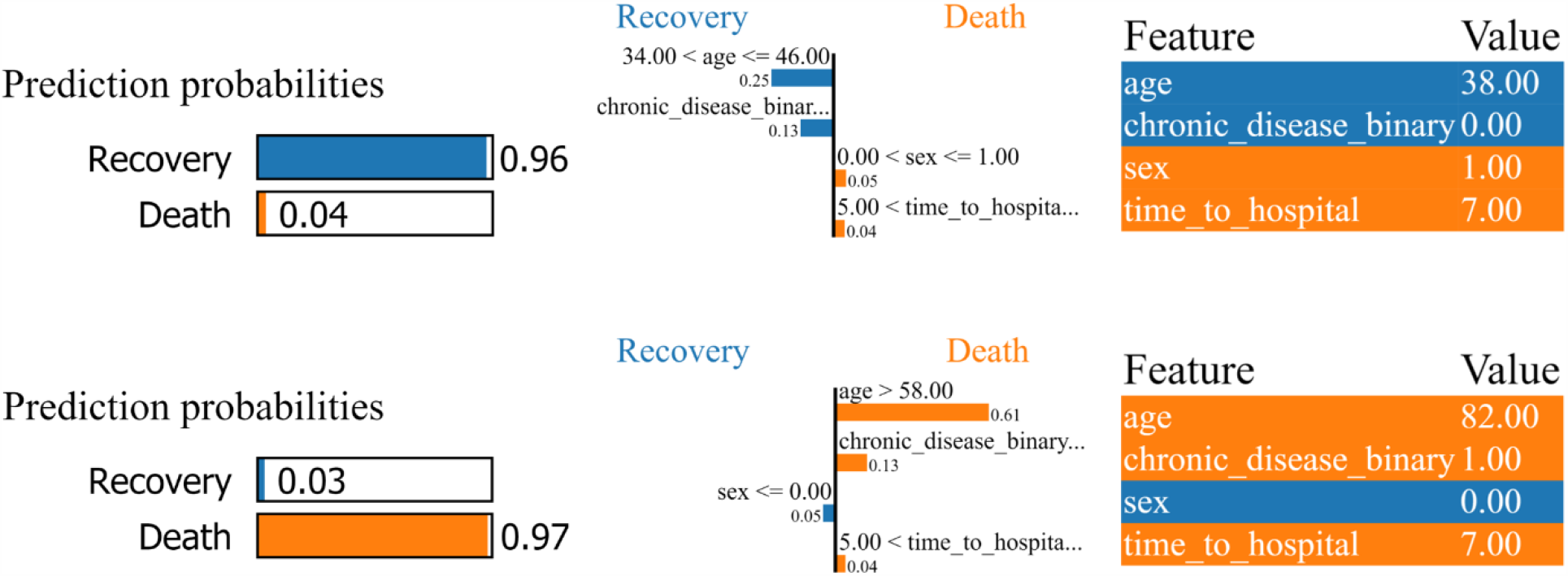
LIME local interpretations for a patient who experienced recovery and was predicted to recover (top) and for a patient who experienced mortality and was predicted to die (bottom).

Skater also provides functionality for creation of partial dependence plots. Fig 9 shows one-way partial dependence plots created by the skater package. These appear to be similar to the plots created using the shap package.

**Fig 9:**
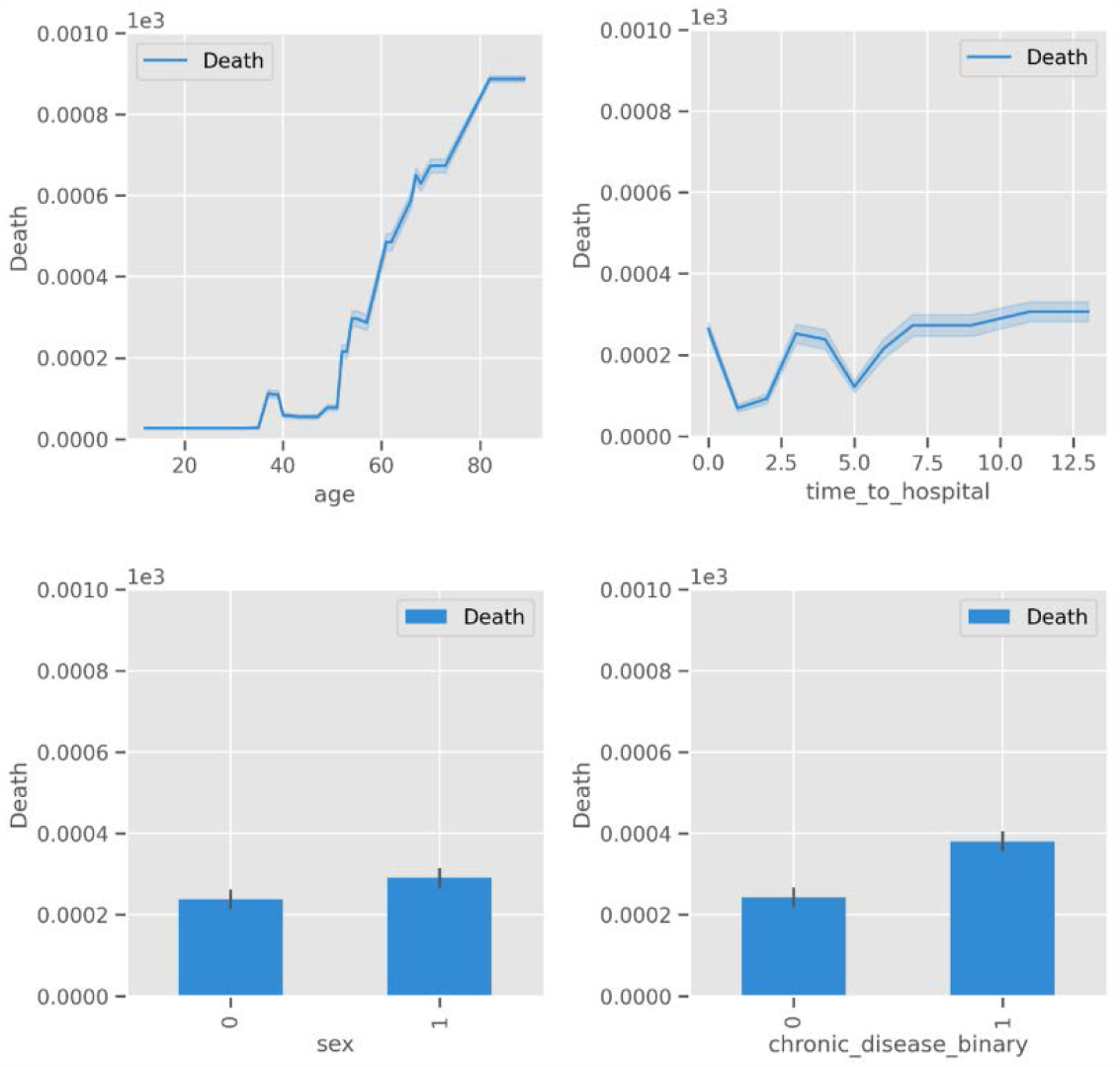
Partial dependence plots with error bars as created by the skater package.

### Surrogate Trees

Although tree-based models are generally considered to be interpretable [20], XGBoost (like other gradient boosting algorithms) combines many trees (100 by default) as weak predictors. More parsimonious trees are required to find simple decision rules (heuristics) for use in a clinical setting. Therefore, we create a parsimonious surrogate tree using the skater package (Fig 10).

**Fig 10:**
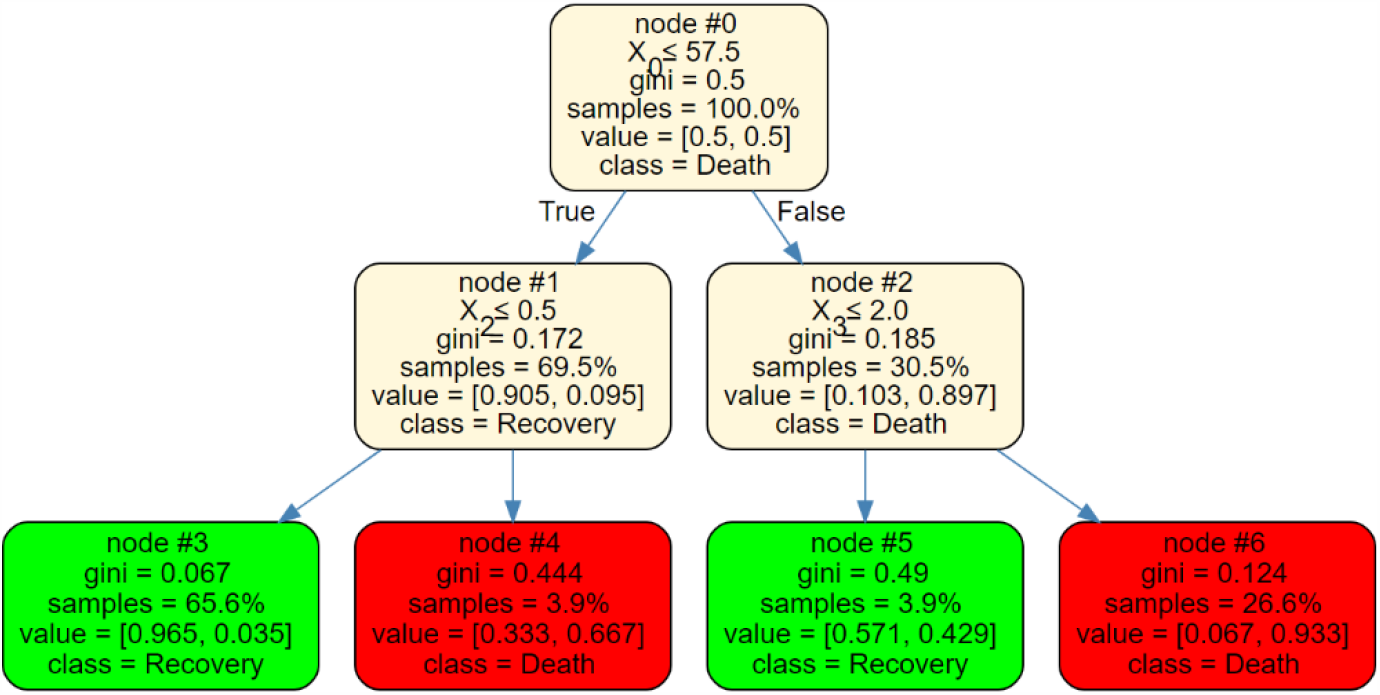
A parsimonious 3-split surrogate decision tree. X0, X1, X2 and X3 are age, sex, chronic_disease_binary, and time_to_hospital respectively.

Rules of thumb can easily be extracted from this parsimonious tree. In this tree, four simple decision rules can be extracted:

1. If the person’s age is 57.5 or less and they do not have chronic disease, the probability of mortality is 3.5%.
2. If the person’s age is 57.5 or less and they have chronic disease, the probability of mortality is 66.7%.
3. If the person’s age is greater than 57.5 and they get to the hospital in 2 days or less (after symptom onset), the probability of mortality is 42.9%.
4. If the person’s age is greater than 57.5 and they get to the hospital after more than 2 days, the probability of mortality is 93.3%.

Note that in this tree, the sex variable was not used, but different trees using different combinations of explanatory variables can be created by tweaking the random seed of the surrogate explainer. Various classification metrics were calculated to assess the prediction performance of the parsimonious model on the test data (Table 3). Interestingly, the more parsimonious model still achieves a classification accuracy of 84% despite only having 3 splits.

**Table 3:** Classification report for 3-split surrogate tree predictions on test set.

|  | Precision | Recall | F1 Score | Support |
| --- | --- | --- | --- | --- |
| **0 (Recovery)** | 0.95 | 0.84 | 0.89 | 44 |
| **1 (Mortality)** | 0.59 | 0.83 | 0.69 | 12 |
| **Accuracy** |  |  | 0.84 | 56 |
| **Macro Avg** | 0.77 | 0.84 | 0.79 | 56 |
| **Weighted Avg** | 0.87 | 0.84 | 0.85 | 56 |

## Discussion

This paper developed an XGBoost model for prediction of individual-level COVID-19 mortality and performed global and local model interpretations using Shapley values from coalitional game theory. Global and local intepretations were also performed using the skater package. Both methods resulted in the similar ranking of the relative importance of the four explanatory variables studied, placing age as the most important feature and time to hospital after symptom onset as the second most important. The interpretation techniques differed in that one ranked sex as more important than chronic disease presence while the other ranked chronic disease presence as more important than sex. Lastly, a surrogate tree model was developed by perturbing the XGBoost model’s inputs and observing the outputs. The surrogate tree achieved a high degree of parsimony while retaining a relatively high predictive accuracy of 84%. Because of its parsimony, the surrogate tree model retains interpretability and can potentially be used in a clinical setting. Furthermore, rules-of-thumb about COVID-19 mortality probabilities can easily be derived by tracing different root-to-leaf paths on the tree.

Hospital systems are not generally well-equipped to handle pandemics, and many hospitals are facing resource shortages. Some estimates suggest that at the peak of the COVID-19 outbreak in the US, the number of ICU beds required would be 3.8 times the number in existence [21]. COVID-19 mortality prediction models can potentially be used to help allocate resources to those with the highest risk of dying in hospitals with limited resources and high load. In addition to developing as a potential tool for clinical resource allocation, this study determines the relative importance of four suspected risk factors and demonstrates the viability of local model interpretations for data-driven clinical decision-making.

To the best of our knowledge, no other published studies have predicted COVID-19 mortality solely off of demographic and temporal variables. This paper demonstrates that COVID-19 mortality prediction can be accomplished with 91% accuracy (or 84% in the parsimonious model) without the use of cellular, molecular, and chemical biomarkers.

Future analysis is required to determine the joint effect of multiple features on outcome and explore other demographic, spatial, temporal, and environmental factors as data on them becomes readily available.

## Data Availability

The original publicly available dataset was taken from this paper: https://www.nature.com/articles/s41597-020-0448-0. The cleaned and filtered data subset of 184 patients used in this paper is available at this repository: https://github.com/yangrussell/covid-19.

## Acknowledgements

None

